# Stress and coping mechanisms among Police Officers in Nigeria: A National Qualitative Study

**DOI:** 10.64898/2026.01.08.26343486

**Authors:** Musibau A. Titiloye, Mojisola M. Oluwasanu, Bibilola Oladeji, Hussain Oluwatobi, Adeyinka Adefolarin, Patrick Okafor, Olayinka Ajayi, Unogu Mackson Osondu, Ezinne Uvere, Ademola J. Ajuwon

## Abstract

Policing is one of the most rewarding occupations; however, it is stressful and demanding. This study was designed to explore stress, stress management, and coping mechanisms among Nigerian Police Officers working across four geopolitical zones in Nigeria.

Using an exploratory design, forty in-depth interviews (IDIs) were conducted with police officers. Data was collected using an interview guide. The interviews were conducted in English and the participants’ indigenous languages (by preference), audio-recorded, and transcribed verbatim. Data were analyzed using the thematic approach.

A range of contextual stressors were identified as barriers to the health and well-being of police officers in Nigeria. The police often lack the tools and equipment needed to perform their official duties effectively. This includes items like uniforms, bulletproof vests, and even operational vehicles. Shortage of manpower, lack of operational tools, poor welfare for police officers, and poor remuneration were also among their concerns. The participants were able to identify signs of stress that are common among police officers, which are majorly weaknesses, lack of sleep, dizziness, headache, anxiety, exhaustion, and anger. The common coping mechanisms include regular exercise, adequate rest, and relaxation through recreational activities, regular medical checkups, and seeking support from colleagues, among others.

Nigerian police officers face many challenges that affect their health and daily routines. This analysis identifies potential opportunities to improve officers’ welfare in these contexts.

## Introduction

Globally, police work is considered one of the most stressful occupations. Police are at high risk of developing work-related stress. Stress is a mental and physical condition that occurs when an individual adjusts to or adapts to the environment, and a perceptual phenomenon arising from a comparison between the demands on the person and his/her ability to cope (1). Stress is associated with increased rates of hypertension, hypercholesterolemia, cigarette usage, higher body mass index among law enforcement workers, depression, domestic violence, and an increase in occupational burnout (2).

Daily exposure to stressful situations is more frequent among police officers than in most other professions (3). The work schedule of a police officer involves long stretches of relative inactivity, unpredictable and stressful bursts of high-intensity events that demand urgent response to life-threatening emergencies (4). Increased work demands impinging on home life, lack of consultation and communication with higher authorities in the organization, lack of control over workload, and inadequate support have been identified as potential factors responsible for stress among policemen (5).

In Nigeria, police officers are exposed to stress from multiple sources, including their occupation, family, and community (6). Police officers often encounter traumatic, critical, and highly dangerous situations, such as violence, abuse, dealing with offenders, accidents, and casualties (7). Their duties include comforting individuals after the loss of a loved one, assisting car accident victims, and listening to assault victims recount their experiences. Due to their work with traumatized individuals(8), Officers may develop symptoms of compassion fatigue, also known as the ‘cost of caring.’ Compassion fatigue is defined as a state of tension and preoccupation with the trauma experienced by others, leading to a desire to avoid the traumatized individuals and any reminders of their trauma, as well as a diminished capacity or interest in bearing others’ suffering (9).

Police officers face multiple challenges, including controlling angry mobs, conducting counter-insurgency operations, managing traffic, and providing security during political rallies and religious festivals, yet they are expected to remain calm and composed while undertaking these tasks (10). Nigeria police officers are vulnerable to many stress-related mental health conditions, including depression, anger disorder, mood swings, burnout, Post-Traumatic Stress Disorders (PTSD), and suicidal ideation (6; 11), risk factors for cardiovascular conditions such as hypertension and abdominal obesity (12).

Studies conducted by (6, 11, 10) confirmed that many police officers in Nigeria face extreme levels of stress, have poor knowledge about stress (11), unhealthy coping mechanisms to deal with occupational stress, including excessive use of alcohol and abuse of stimulants (10), and that they deploy excessive aggression on the citizenry (13). Yet, given the important role they play in maintaining law and order, police officers need to be mentally and emotionally healthy to perform their duties effectively and efficiently.

A study by (10) showed that the majority (95.9%) of police personnel were engaged in field duties, with only 4.08% assigned to office tasks. Among these constables, several factors contributed to significant psychological stress, including inadequate housing and family security, irregular working hours, accountability, and transfers, lack of personal time, insufficient recognition for good work, overtime work, inadequate salary, lack of holidays, limited opportunities for advancement, and delayed promotions (14). As the first responders to many risky situations, police officers need skills to successfully manage stress not only for their mental health but also for the safety of the communities they serve (15). The main objective of the study reported in this article was to assess the burden of stress, its management, coping mechanisms, and related mental health conditions among police officers in Nigeria using a qualitative approach. The collection of qualitative data is expected to provide insights as part of the first steps to developing appropriate interventions to address this problem among police officers in Nigeria.

## Methodology

### Study Design

The study is exploratory in design. It is a component of a large study that investigated the burden of stress among police officers in Nigeria. The study is qualitative, and in-depth interviews (IDIs) were conducted to document personal experiences, coping mechanisms, stress management, and related mental health issues. The adoption of a qualitative approach aimed to collect data that would provide insights into the burden of stress and how this problem can be addressed among police officers in Nigeria.

### Study Sites

A simple random sampling technique (balloting) was used to select four states, namely Bauchi, Nasarawa, Akwa-Ibom, and Oyo, selected from four(North-East, North-Central, South-South, and South-West) out of the six geo-political zones of the country in which the study was conducted.

### Study Population

The study population comprised male and female police officers at the study sites. Both senior and junior officers were selected. The Principal Investigator/research team conducted advocacy visits to the Police Headquarters in Abuja and to each of the State Police Commands to obtain official permission necessary to facilitate the smooth entry and participation of the police community in the study. Upon approval, contact was made with either the state Commissioner of Police or the State Police Medical Officer, who issued directives to all selected divisions in each state to commence the study. A liaison police officer in each state worked with each team lead to facilitate access to potential respondents.

### Instrument for Data Collection

An in-depth interview (IDI) guide was developed and used for data collection. The guide consisted of thirteen (13) point questions to elicit demographic information about the respondents, personal experiences with occupational and organizational stressors, coping and management mechanisms to deal with stress-related issues. The questions were framed in English, the country’s official language.

### Recruitment of study participants

With an estimation of sixty (60) police divisions in each state, about thirty (30) of the divisions in each state were randomly selected to cut across the three senatorial districts, reflecting the potential diversity in each state. Ten (10) Police officers were interviewed in each state. Efforts were made to select 10 divisions per senatorial district in each state. In each division, a list of all police officers was obtained from the divisional police officer, and 10 officers were purposively selected. The criteria for selecting participants were willingness and consent to participate in the study, a schedule that involves direct interactions with community members, attachment to a defined police formation, and availability throughout the period of the research work.

### Recruitment Research Assistants

Ten research assistants (RAs) and two supervisors were recruited locally in each state (a total of 48) and trained to collect data. Training focused on the research objectives, interview procedures, interpersonal communication, and ethical issues. The RAs were trained to conduct the interviews under the supervision of trained supervisors, while a member of the investigating team was assigned to a state for field coordination and monitoring.

### Procedure for data collection

The interviews were conducted between the 9^th^ and 20^th^ of August, 2023. All interviews were conducted at a convenient place within the police office premises. The RA provided detailed information on the study’s objectives to each participant, and written consent was obtained from each interviewee before the commencement of each interview. The interviewer informed each prospective participant that participation in the study was voluntary and clarified any questions the participant had about the issues discussed in the guide. In all, forty (40) IDIs were done in the four sites, ten (10) in each state. All interviews were conducted at a time that was convenient for the participants.

### Data Collection Procedures

Privacy was maintained throughout the interview. All interviews were recorded on a digital recorder after consent had been obtained from each study participant. Each interview lasted 50 to 60 minutes.

#### Data Analysis

The data were transcribed verbatim, which enabled analysis using the thematic method. Verbatim transcription was performed close to the completion of the interviews/discussions by the interviewers to maintain the originality of the information without loss of themes. Data transcription was performed under the supervision of the team leader, who reviewed it for completeness. The data analysis process commenced with the development of a codebook by the research team to guide coding and ensure consistent classification of themes. Notes taken during the interview were also consulted to ensure the accuracy of the information and to support quality assurance strategies. Thematic analysis was used to code the documents and transcripts. The Consolidated Criteria for Reporting Qualitative Research (COREQ) (16) was adopted for presenting and reporting the data.

#### Quality assurance

To ensure the collection of high-quality data, data transcription occurred in parallel with data collection and was shared on an ongoing basis with the study team leader to maintain data quality. State team leaders were in constant communication with the interviewers in the field to address any issues during the data collection period. Adequate training was conducted to capacitate all the field staff through a well-developed training manual.

## Ethical Considerations

The National Health and Research Ethics Committee (NHREC), Federal Ministry of Health, Abuja (NHREC/01/01/2007-25/04/2023) approved the protocol before the commencement of data collection. The Inspector General of Police and the Commissioner of Police in each state also granted permission for the study. All participants were duly informed of all the procedures involved and that participation was voluntary. Written consent was sought before recruiting them for the study.

## Findings

### Demographic Information of Participants

The majority of the police officers who participated in this qualitative component of the study are males (87.8%), while more than half (51.2%)were between the 41-50yearsage bracket. Almost three-quarters (75.6%) are Christian. Almost half (48.8%) have spent more than 20 years in the Nigerian Police Force; 75.6% have more than a Senior Secondary School Certificate; more than half (63.4%) are Senior officers; and

**Table 1:** Demographic Information of Participants.

| **Demographic Information** | **States** | **States** | **States** | **States** |
| --- | --- | --- | --- | --- |
| **Demographic Information** | **Akwa Ibom(%)** | **Bauchi (%)** | **Nasarawa (%)** | **Oyo (%)** |
| **Sex** |  |  |  |  |
| Male | 9 (90.0) | 11(100.0) | 10(100) | 6(60.0) |
| Female | 1 (10.0) | 0(0.0) | 0(0.0) | 4(40.0) |
| **Age:** |  |  |  |  |
| 21-30 | 1 (10.0) | 0(0.0) | 2(20) | 1(10.0) |
| 31-40 | 2 (20.0) | 2(8.2) | 2(20) | 2(20.0) |
| 41-50 | 4(40.0) | 9(81.8) | 4(40) | 4(40.0) |
| 51-60 | 3 (30.0) | 0(0.0) | 2(20) | 3 (30. |
| **Religion** |  |  |  |  |
| Islam | 0(0.0) | 8(72.7) | 1(10.0) | 1(10.0) |
| Christianity | 10 (100.0) | 3(27.3) | 9(90.0) | 9(90.0) |
| **Number of years in service** |  |  |  |  |
| ≤10 | 2 (20.0) | 0(0.0) | 3(30.0) | 2(20.0) |
| >10-20 | 2 (20.0) | 8(72.7) | 2(20.0) | 2 (20. |
| >20-30 | 4 (40.0) | 3(27.3) | 5(50.0) | 4 (40. |
| >30-40 | 2 (20.0) | 0(0.0) | 0(0.0) | 2(20.0) |
| **Highest level of Education** |  |  |  |  |
| SSCE | 2 (20.0) | 3(27.3) | 4(40.0) | 1(10.0) |
| University degree | 8 (80.0) | 8(72.7) | 6(60.0) | 9(90.0) |
| **Rank** |  |  |  |  |
| **Senior Officer** | 6(60.0) | 6(54.5) | 6(60.0) | 8(80.0) |
| **Junior Officer** | 4(40.0) | 5(45.5) | 4(40.0) | 2(20.0) |
| **Marital Status** |  |  |  |  |
| Single | 0(0.0) | 1(9.1) | 1(10.0) | 0(0.0) |
| Married | 10(100.0) | 10(90.9) | 8(80.0) | 10(100 |
| Divorced | 0(0.0) | 0(0.0) | 1(10.0) | 0(0.0) |
| **Year since on the same rank** |  |  |  |  |
| 0-5 | 8(80.0) | 8(72.7) | 8(80.0) | 3(30.0) |
| >5-10 | 2(20.0) | 3(27.3) | 1(10.0) | 7(70.0) |
| >10 | 0(0.0) | 0(0.0) | 1(10.0) | 0(0.0) |

#### General concerns and challenges about the police workforce

Respondents stated that the primary responsibilities of the police force include combating and preventing crime, investigating criminal activities, protecting the community, and providing security for political officeholders. Several concerns and challenges were identified, including low salaries, a lack of tools, equipment, and manpower.

Poor remuneration is a major concern, as it can demotivate officers and make it difficult for them to support themselves and their families. Furthermore, police officers often lack the necessary tools and equipment to effectively carry out their duties. This includes items like uniforms, bulletproof vests, and even operational vehicles. These concerns are illustrated in the quotes below

> *“The number one biggest challenge of the police force, I will say, is our remuneration. I mean, the salary is very low. And the number two is we don’t have tools, and when I say tools, I mean tools to work with” (35-40 years old Police Officer, Northern Region).*

> *“The challenges we are having are numerous, including a lack of manpower and a lack of equipment. Secondly, is that financially, with the current situation of Nigeria, we cannot fuel this car to maximally use for security purposes (50-55 years old Police Officer, Northern Region)”.*

The nature of the work means officers can be called upon at any time to respond to emergencies. Police officers are essentially on a 24-hour duty, except during specific periods when there are no immediate issues. This constant readiness is a fundamental aspect of police work, and officers don’t have fixed rest periods. Officers often find themselves handling the workload of multiple individuals, further increasing their stress and workload. These challenges are compounded by the insufficient personnel, making it difficult to effectively address the security needs of the population, resulting in extended work shifts with resultant effects on their health and stress levels as expressed in the quotes below:

> *“The police force generally is a very stressful zone to work. They have three shifts: morning shifts, afternoon shifts, and evening shifts, but now you rarely have three shifts; you have two shifts. (50-55 years old Police Officer, Southern Region)”.*

> *“We are short of manpower, and that is why we face the challenge of even doubling our duty. We are being scheduled for 8 hours or more because we do not have enough manpower. The major problem is manpower. (50-55 years old Police Officer, Southern Region)”.*

> *“……… as a police officer, we have numerous challenges and our challenges are basically, Manpower challenge…… (40-45 years old Police Officer, Northern Region)”.*

Police officers often face significant difficulties meeting their basic needs and maintaining their well-being, and many struggle to find suitable housing, making it challenging to establish a stable living arrangement. Police officers posted in areas with security challenges often have difficulty accessing medical care. The lack of access to healthcare can lead to delays in treatment and higher costs for officers who need medical care. These challenges are illustrated in the quotes below:

> *“Then I used to go to that clinic every month, complaining to them that I’m sick. They would tell me to go and do tests. The one that you are supposed to buy and pay a little money, they refer you to get it outside (45-50 years old Police Officer, Southern Region)”.*

> *“And there is no fund meant for the police for such, to take immediate treatment. You end up struggling, borrowing, and contributing voluntarily to take care of the person. These are the challenges that we have. (55-60 years old Police Officer, Southern Region)”.*

#### Knowledge about stress, causes, signs, and symptoms

Participants expressed different notions of the meaning of stress. According to them, stress is described as a state of being overwhelmed by activities, leading to physical and mental breakdowns. Participants highlight the impact of long working hours, insufficient rest, and a challenging work environment, especially in a profession that operates around the clock with no holidays or breaks. Stress manifests as fatigue, anxiety, and disrupted sleep patterns.

The interviewees link stress to external factors such as financial problems, family issues, and the overall work environment. They emphasize the importance of welfare and support systems in mitigating stress, emphasizing that when individuals feel taken care of, stress is reduced. Overall, stress is perceived as a result of factors beyond an individual’s control, leading to a state of mental instability and physical fatigue. Participants across the states gave several definitions of stress as illustrated in the quotes below:

> *“Stress is a state of being overwhelmed with activities that break down your system. The causes of stress are numerous. Some could be biological, some environmental. Like most of the police, it is an environmental condition. The signs and symptoms of stress are headache, weakness of the body, muscle cramp (55-60 years old Police Officer, Southern Region).”*

> *“Stress can be defined as mental instability after a long time at work. Physical and mental inability to do other things, and fatigue after a while. Not relaxing after work after a long period of being active, and for some people, financial constraints”. (25-30 year olds Police Officer, Southern Region).*

> *“Always getting tired, heart beats a lot, anxiety, mental tiredness. Lack of equipment, long hours in performing the job. Attitudes toward work, no motivations, fatigue, walking slowly, and easily provoked on just a small task” (45-50 year olds Police Officer, Northern Region)”.*

### Health issues related to organizational stress

The majority of the police officers interviewed said the health issues related to organizational stress include not having enough time to rest, several stated that they experience headaches, malaria, typhoid, dizziness, and weakness in the body. Some have hypertension and insomnia, cold and pneumonia, and when this happens, there is a loss of focus and concentration. Several health hazards were buttressed by the quotes below:

> *“We experience headaches, blood pressure issues, in fact, all” (45-50 years old Police Officer, Southern Region).*

> *“………. I work 16 hours, and I come back home, and I cannot sleep again, umm, loss of appetite, I get a lot of cramps, physical cramps, oh when I am stressed I will go through 30 minutes of muscle pull because I have not rested” (40-45 years old Police Officer, Northern Region).*

> *“The health issues as a result of stress are arthritis, stroke, and high blood pressure. Those are the major ones, headache, malaria, and fever” (45.50 years old Police Officer, Southern Region)*

There is also ill-health caused by bullet wounds, harsh weather, tear gas, and hazards on the road, as illustrated in the quote below:

> *“Tear gas is very harmful. No matter how you shoot it, you must inhale it. So… we are fallible, we make mistakes, at times you hear something like accidental discharge, or he shoots a colleague, or he shoots himself, in operations, you have a lot of health challenges” (45-50 years old Police Officer, Northern Region).*

### Perception about policing

Policing has a negative multiplier effect on respondents’ work, though they are trained to always persevere. Few stated that they perceive their work as not tiring, as it is a job they could do for 35 years. The respondents’ overall perception of their job being positive, is illustrated in the quotes below:

> *“By the grace of God, I don’t feel like I’m tired of the work because it’s a job of 35 years” (45-50 years old Police Officer, Southern Region)*

> *“It’s a good one, I love it, I love it. Doing all this, I have to say, I love my job, in the sense that I have my confidence, that am saving my own country, I don’t think I will have a job I will love like the police job, because people are looking up you day by day and by the grace of God, I will not fail them” (45-50 years old Police Officer, Northern Region).*

> *“I perceive this work as the most important to me. I like it. And I do it with passion. I am passionate about this work, and I take great delight in it. Despite the stress, I enjoy doing the work because it gives me many chances to relate with people” (55-60 years old Police Officer, Southern Region).*

### Coping strategies and management of stress

Interviewees reported that police officers adopt different coping mechanisms to manage stress. Some of these are taking casual leave to sleep and rest; depending on God, some rely on their spouses and family members. They often seek psychological and medical help, take medicines, or practice relaxation techniques such as yoga, deep breathing, and watching football. Also, being patient during a stressful situation, listening to music, self-care, and reading books were coping mechanisms adopted as corroborated by the participants below:

> *“I’m getting at least off for maybe a few hours, and if it has been granted unto me, then I will go and rest” (used to it because the only way I can manage it, maybe after work, I will have few hours of rest, no matter what, in order to bring down the level of stress in me. Once in a while, I write a leave permit (30-35 years old years old Police Officer, Northern Region).*

> *“I apply for casual leave, which will give me room to rest enough so that I can manage that stress” (40-45 years old Police Officer, Southern Region).*

> *“Coping with stress is something I have learnt to do. At times, I watch movies, read books when I’m not busy. I sleep most of the time. I take time to rest, eat, and sleep (25-30 years old Police Officer, Southern Region)*

### Suggested interventions to address stress

Participants suggested six different interventions. These included compulsory routine medical check-ups, training programmes to address stress, good salary packages, a conducive working environment, such as operational vehicles to carry out their duties efficiently, and the recruitment of more junior officers to address the shortage of manpower. The policemen also suggested that the government provide modern equipment to aid their work. Their suggestions are captured in the quotes below:

> *“recruitment of additional manpower ……… When there is sufficient manpower, those doing the work will be very happy and have time to rest; there may be no overtime or lack of motivation. The government should provide us with appropriate medication for us to cope with the high cost of treatment of stress” (40-45 years old Police Officer, Northern Region).*

> *“Now imagine we have drones in my unit as anti-kidnapping; no matter where a human being is, we can pick them up, and we just go there to rescue the victims. We spend days and hours without succeeding, which is very stressful. If police personnel could have access to a very good health facility for normal and regular medical checks, that would reduce stress” (40-45 years old, Police Officer, Northern Region).*

> *“What I am suggesting now is we should encourage more of this type of research that will bring measurable results that can be implemented. Over the 13 years I have been on this job, this is the first time I am meeting any organization or anybody talking about stress, and it is our major performance.” (40-45 years old Police Officer, Northern Region).*

It was also suggested that the government should address the manpower shortage by recruiting more police officers, so that those in the system will be less stressed. The government should address the inadequate accommodation by building more police barracks to accommodate all police personnel, as other armed forces do. Police officers living in the barracks will provide more security consciousness than those living outside the barracks; a condition in which some of the officers in Nigeria have found themselves, as illustrated in the quote given below:

> *“About recruitment to the police, before anybody is recruited into the police force, they should go through a psychological test to make sure that the person does not have any mental health-related issues” (55-60 years old Police Officer, Southern Region).*

> ***“**My suggestion is that the government should put in place things that will make the police work effectively. And they should do it in a way that it will meet the end user, so it cannot be lost along the way” (45-50 years old Police Officer, Northern Region).*

> *“Allowances for special operations and duties are not being paid to the respective officers assigned. These have put a lot of pressure on their finances because they have to borrow money from others to fend for themselves (40-45 years old Police Officer, Southern Region).*

## Discussion

Findings from our study revealed that participants had some level of awareness of stress, which is in line with the study by John-Akinola et al. (2022), which showed that police officers had poor knowledge of stress. They were able to identify stress-related symptoms, including some health conditions associated with stress. Participants in this study were particularly aware of several contributory factors to stress, including external factors such as financial problems, family issues, and insufficient welfare packages. These findings aligned with those studies that identified similar factors, such as inadequate salary level, shift work, and family-work conflict as causes of stress (6).

Two types of coping mechanisms, positive or negative, emerged from the interviewees. The positive coping mechanisms adopted by participants in this study included taking casual leave in other to rest, engaging in some form of physical exercises (yoga, deep breathing), having social relationships with spouses and family members; seeking psychological counseling and medical help (including taking medicine) and other leisure activities (including watching football, reading books, listen to music). The above positive mechanisms are similar to those reported in the literature (17, 18, 19).

Nigeria is a predominantly religious country with two dominant religious groups – Christians and Muslims. Religion appears to be a widely accepted means of solving a variety of human problems, including stress (19). Reliance on religious belief emerged as a major mechanism for coping with stress. It has been documented that greater involvement in religious activities increases the likelihood of adopting preventive health behaviors (20). Evidence from the literature further supports religiosity as a strategy to cope with stress (19). While this approach offers some benefits and relief, it can be inferred as a defensive strategy.

Findings from this study reveal the presence of both operational and occupational stress, along with associated stressors. The categorization of stress into these two groups has been established in the literature (13). Some of the operational stressors included the lack of necessary work tools, equipment, operational vehicles, and access to financial support towards the purchase of or replacement of uniforms. Should these stressful situations continue unaddressed, the quality of decisions made by the police officer could be reduced, thereby increasing anger and aggressive tendencies (13). A state of constant irritation can cause a Police officer to react negatively and aggressively to even the slightest provocation, misinterpreting the event’s magnitude or seriousness as potentially harmful (13).

Organizational stress identified during this interview included a lack of adequate manpower, re-training opportunities for staff, low welfare schemes, remuneration challenges, excessive work shifts, work conflicts, and the absence of equal opportunity in decision-making concerning issues that bother them. These findings are further corroborated by a previous study (17).

Previous studies have documented the presence of these organizational stressors among Nigerian police units (21) and have further revealed that organizational stressors create greater stress for Police officers than operational stressors. This further heightens the impact of stress, associated morbidities, and psychosocial vices (21), including an increase in high blood pressure rates, elevated risk of cardiovascular diseases, insomnia, increased levels of destructive stress hormone, heart problems, post-traumatic stress disorder, suicide, and family instabilities, among others.

The identification of both stressors, which cut across many professional careers, especially the police force, is important given their negative implications for staff performance, staff health, organizational efficiency, viability, and sustainability in the long run (21). Hence, the utmost priority should be placed on revising and creating both existing and work-friendly policies to address specific stressors that can lower police staff performance.

### Strengths/Limitations of this study

To the best of our knowledge, this is the first national study that spans four of the country’s six geopolitical zones. As a national study, the findings provide deep-seated insight into the problems and stress-related issues that Nigerian police officers are bedeviled by.

The views expressed were first-hand information from the respondents, which provides the evidence required to provide context-specific and sustainable interventions to address the issues at hand. The only limitation is that the findings are not generalizable, as they were obtained qualitatively.

## Conclusion

There is a dire need to create an enabling environment (in the form of work-friendly policies), increase salaries, wages, and other emoluments, improve the accessibility and affordability of medical services, and provide appropriate and adequate manpower to encourage and assist police officers in stress prevention. This is important because reliance on defensive coping strategies in the absence of addressing the underlying factors contributing to the stress may offer transient relief, which, in themselves, are insufficient given the multiple challenges confronting the Nigerian state, which the police are expected to address in the interest of national peace and security.

## Data Availability

Transcripts can be provided upon request to the corresponding author. This can only be used for non-commercial purposes which ensures that participants’ confidentiality is protected

## Acknowledgments

The authors wish to acknowledge the study participants for participating in the study and the Nigeria Police Force for their involvement.

## Notes

### Competing Interest Statement

The authors have declared no competing interest.

### Funding Statement

The study is funded by the Tertiary Education Trust Fund (TETFund), Nigeria, under the National Research Fund Scheme (TETF/DR&DCE/NRF2021/SETI/ HSW/00183/-01/VOL.1). The funders have no role in the design of the study collection, management, analysis, and interpretation of data writing of the report and the decision to submit the report for publication.

### Author Declarations

The National Health and Research Ethics Committee (NHREC) of the Federal Ministry of Health, Abuja, Nigeria approved the protocol (NHREC/01/01/2007-25/04/2023) before the commencement of data collection.

## References

1. Coon D, Mitterer JO. Introduction to Psychology: Gateways to Mind and Behavior,.. Study Guide. Wadsworth Cengage Learning; 2010.

2. Ogungbamila B, Fajemirokun I. Job stress and police burnout: Moderating roles of gender and marital status. IAFOR Journal of Psychology and the Behavioural Sciences. 2016;2(3):17–32.

3. Purba A, Demou E. The relationship between organisational stressors and mental wellbeing within police officers: a systematic review. BMC Public Health. 2019 Dec; 19:1–21.

4. Kales SN, Soteriades ES, Christophi CA, Christiani DC. Emergency duties and deaths from heart disease among firefighters in the United States. New England Journal of Medicine. 2007 Mar 22; 356(12):1207–15.

5. Johnson O, Jaeckle T. Mitigating the emotional impact of stress on law enforcement personnel. Bureau of Justice Assistance, US Department of Justice. 2018 Apr 1.

6. Lateef A. Exploring the factors responsible for occupational stress among police officers in Nigeria (Doctoral dissertation, Walden University

7. Queirós C, Passos F, Bártolo A, Marques AJ, Da Silva CF, Pereira A. Burnout and stress measurement in police officers: Literature review and a study with the operational police stress questionnaire. Frontiers in Psychology. 2020 May 7; 11:587.

8. Costa T, Passos F, Queiros C. Suicides of male Portuguese police officers–10 years of national data. Crisis. 2019 Jan 15.

9. Ondrejková N, Halamová J. Stressful factors, experiences of compassion fatigue and self-care strategies in police officers. Journal of Police and Criminal Psychology. 2022 Dec; 37(4):892–903.

10. Ruwani II, Shinkut NP, Ishaya DS, Iliya V. Occupational stress and coping strategies among the Nigeria Police Force in Kaduna state. International Journal of Management, Social Sciences, Peace and Conflict Studies (IJMSSPCS). 2020;3(3):77–96.

11. John-Akinola YO, Ajayi AO, Oluwasanu MM. Experience of stress and coping mechanisms among police officers in south western Nigeria. International Quarterly of Community Health Education. 2020 Oct; 41(1):7–14.

12. Hussain OJ, Ajuwon AJ. Prevalence, Knowledge and preventive practices against Hypertension among police officers in Ibadan. Annals of Ibadan Postgraduate Medicine. 2020;18(2):114–21.

13. Akindele-Oscar Y, Bello SO, Aborishade R. The Influence of Occupational Stress and Mental Health on Nigerian Police Officers™’ Aggression. NIU Journal of Social Sciences. 2019 Oct 23;5(3):165–70.

14. Indicators OE, Hagvísar OE. Health at a glance 2019: OECD indicators. Paris: OECD Publishing; 2019.

15. Patterson GT, Chung IW, Swan PG. The effects of stress management interventions among police officers and recruits. Campbell Systematic Reviews. 2012; 8(1):1–54

16. Tong A, Sainsbury P, Craig J. Consolidated criteria for reporting qualitative research (COREQ): a 32-item checklist for interviews and focus groups. International journal for quality in health care. 2007 Dec 1;19(6):349–57.

17. Gershon RR, Lin S, Li X. Work stress in aging police officers. Journal of occupational and environmental medicine. 2002 Feb 1;44(2):160–7.

18. Udeh OS, Aguwa EN, Onwasigwe CN. Perceived workplace stress levels and coping strategies of military personnel in a Nigerian barrack. Journal of Community Medicine and Primary Health Care. 2022 Dec 8;34(3):110–25.

19. Wakil AA. Occupational stress among Nigerian police officers: An examination of the coping strategies and the consequences. African Research Review. 2015 Oct 27; 9(4):16–26.

20. Idehen EE, Kehinde OA. Religiosity and the preventive health behaviour of young adults. IFE PsychologIA: An International Journal. 2010 Mar 1; 18(1):183–8.

21. Adegoke TG. Effects of occupational stress on psychological well-being of police employees in Ibadan metropolis, Nigeria. African Research Review. 2014 Feb 21; 8(1):302–20.

